# Association between virtual visits and health outcomes of people living with HIV: A cross-sectional study

**DOI:** 10.1101/2024.12.04.24318511

**Authors:** Nadia Rehman, Lawrence Mbuagbaw, Dominik Mertz, Giulia M. Muraca, Ontario HIV Treatment Network Cohort Study, Realize, Aaron Jones

## Abstract

**Background:** Virtual care has been integrated as a modality of care in Ontario, yet its effectiveness for people living with HIV remains largely unexplored.

**Objectives:** We aimed to determine the association of visit modality (virtual, in-person, or both) on adherence to antiretroviral therapy (ART), viral load, and quality of life (QOL) in people living with HIV in Ontario, Canada.

**Methods:** We conducted a cross-sectional study using data from the 2022 Ontario HIV Treatment Network Cohort Study (OCS), collected during the COVID-19 pandemic when virtual visits were first introduced. Participants were grouped into three categories based on the mode of care: virtual, in-person, or a combination of both. Data were collected through self-reported questionnaires and medical records, with viral load data linked to Public Health Ontario Laboratories (PHOL). Logistic regression was used to examine the outcomes of optimal ART adherence and viral load suppression, and linear regression was used for quality of life (mental and physical) outcomes.

**Results:** In 2022, 1930 participants accessed HIV care in the OCS. Among them, 19.0% received virtual care, 45.6% received in-person care, and 34.3% received care through virtual and in-person modalities. The median age of the participants was 55 years (IQR: 45-62). In the multivariable logistic regression model, virtual care was associated with an increased likelihood of optimal adherence to antiretroviral therapy (Adjusted Odds Ratio (AOR) 1.30, 95% confidence interval (CI): 1.00-1.70) and an increased likelihood of achieving viral load suppression (AOR 1.67, 95% CI:1.03-2.63). Moreover, combined virtual and in-person care is associated with an improved mental quality of life compared to in-person care (Adjusted Mean difference (MD) - 0.960, 95% CI 0.052,1.869).

**Conclusion:** This study suggests virtual care is positively associated with adherence to antiretroviral therapy (ART) and viral suppression within this context. However, future research is necessary to establish causality and to assess the long-term effects of virtual care.

## Introduction

HIV is a chronic health challenge, with 22,461 people living with HIV in Ontario, Canada, as of 2020 [1] This population faces a disproportionate burden due to socio-economic disparities and structural barriers within the patient-provider relationship, which further complicates the continuity of HIV care, contributing to sub-optimal retention in healthcare services and poor adherence to antiretroviral therapy (ART) [2]. This results in detectable viral loads, increased opportunistic infections, and higher morbidity and mortality rates [3-6]. Consistent access to ART and regular health assessments necessitates strong patient retention in care, as highlighted by several studies [7-9]. Despite universal public coverage in Ontario, challenges with retaining individuals in care persist. A fully accessible, patient-centred healthcare system is essential to addressing these issues [10].

In 2021, Ontario, Canada, adopted virtual care in the healthcare sector to help mitigate the impact of the SARS-CoV-2 pandemic [12]. Virtual care is a health care model in which all clinical interactions between the practitioner and the patient are delivered using electronic mediums, such as video conferencing or audio digital tools, such as telephone [11]. Since 2022, Ontario has taken concrete measures to integrate virtual visits as complementary to traditional care. These measures include enhancing data security and privacy, addressing social and ethical considerations, establishing regulations for virtual healthcare, and introducing fee codes that enable physicians to bill for virtual visits [10, 12].

Virtual visits can help overcome barriers in HIV care by offering convenience, accessibility, and affordability, potentially improving access to care and patient retention [13, 14]. However, virtual care has several limitations, such as the inability to assess clinical conditions objectively, the lack of necessary equipment, and the absence of physical interactions with physicians, which can lead to mistrust, multiple appointments, and potential misdiagnosis [13]. Technology can also pose barriers for certain groups, including older adults, individuals with lower literacy levels, and those from disadvantaged socio-economic backgrounds [14]. The choice of virtual care compared to in-person visits may be influenced by physician preferences and the significant discrepancies in Ontario’s fee codes between comprehensive versus limited care and telephone calls versus video visits [10, 15]. While some data suggest virtual care improves retention in care, others report increased loss to follow-up [16]. As virtual care evolves, it is essential to address disparities, prevent overuse, and consider unintended financial consequences with health equity and ethical considerations in mind [10, 11, 16-18].

While virtual care has become a standard practice in Ontario [10], the evidence of its effectiveness remains limited. We developed this study alongside the Ontario HIV Treatment Network (OHTN) [19], utilizing the data from the Ontario HIV Treatment Network Cohort Study (OCS), the largest community-governed HIV cohort in North America [20]. The primary objective of the study was to assess whether there were differences in adherence to antiretroviral therapy (ART), quality of life (QOL), and viral load among people living with HIV in Ontario, Canada, based on whether they used virtual or in-person appointments with an HIV care physician. The secondary objective of the study was to evaluate the differences in health outcomes (adherence to ART, QOL, and viral load) among people living with HIV from various socio-demographic and health-related factors in Ontario, as virtual care may affect certain groups differently.

## Methods

### Study Design

We conducted a cross-sectional study using data from participants in the OCS in 2022.

#### Setting

The OCS is a multi-site clinical cohort of people receiving HIV care in Ontario, Canada’s most populous province (population: 13.6 million). Recruitment occurs at ten participating sites, including outpatient clinics in hospitals and community-based practices. The cohort has been described elsewhere [21].

#### Stakeholder Engagement

To achieve our study objectives, we established a community advisory board (CAB) in collaboration with [Realize](https://www.realizecanada.org/), a Canadian charitable organization working with people living with HIV and related organizations. The CAB comprises representatives from various key populations of people living with HIV, enhancing the external validity of our project, bolstering individual and community capacity, and ensuring the effective implementation of our research findings [22, 23]. We convened a meeting with the CAB to seek their insights on the relevance of the research question, the socio-demographic factors involved, and the correlation with the health conditions of people living with HIV. The CAB also actively interpreted the findings and collaborated on a dissemination plan for our research outcomes.

#### Data Sources

Clinical data are collected during routine follow-up visits, sourced from clinic records via manual chart abstractions or computerized medical record systems, and record linkage with Public Health Ontario Laboratories (PHOL), the sole provider of such testing provincially. Additionally, annual interviews are conducted using standardized questionnaires to gather socio-demographic and psycho-social-behavioural information. All participants provide written informed consent, and the cohort design and consent forms are approved by the University of Toronto Research Ethics Boards (REBs) and REBs at each participating site [21]. Data was accessed on November 23, 2023. And the data was in de-identified form.

#### Population

Eligible participants for our study are those aged 16 years or older who visited their HIV physician using any of the three modalities of care (virtual, in-person visits, or both virtual and in-person care) in 2022 and completed the OCS questionnaire. Participants with incomplete information on the type of care received were excluded.

### Measures/Outcomes

#### Primary outcome

Adherence to ART was assessed using self-reported data from the standardized OCS questionnaire. For this study, we aimed to dichotomize adherence into optimal adherence (≥95%) and suboptimal adherence (<95%). We considered optimal adherence as participants who reported never missing a dose or missing a dose more than three months ago. In contrast, suboptimal adherence included those who missed doses within the past week, 1–4 weeks ago, 1-3 months, or responded with "don’t know." These classifications were established in consultation with HIV specialists from a dedicated HIV care facility.

#### Secondary Outcomes

Viral load suppression was defined as ≤ 40 copies/mL, indicating virological suppression (stable viral load). Any value > 40 copies/mL is considered virologically unsuppressed and unfavourable. QOL was assessed using the Short Form 12-item Health Survey (version 2), which includes two components: the Mental Component Summary Score (MCS) and the Physical Component Summary Score (PCS), both of which are reported separately [24].

### Variables

#### Primary exposure

We categorized the HIV care that the patients received into three mutually exclusive categories: i). In-person care at the clinic ii). Virtual care either by telephone or video call, iii). Combination of in-person and virtual forms of care.

#### Demographic and clinical variables

Baseline demographic and clinical data were extracted from the 2022 OCS questionnaire. Due to multiple categories in the OCS data, some of which were not information-rich, we consolidated them into meaningful categories for our study. Data missing values are reported separately in Table 1, which details the baseline characteristics.

**Table 1.** Comparison of baseline characteristics between participants who accessed HIV care through virtual, in-person or both virtual and in-person care in Ontario, Canada, in 2022.

| Characteristics | In-person care<br>n (%) | Virtual care<br>n (%) | Virtual and in-person care<br>n (%) | p-value |
| --- | --- | --- | --- | --- |
| No. of participants | 900 | 367 | 663 |  |
| *Median age [Q1-Q3]* | 55 [44-63] | 56 [46-62] | 55 [45-62] | 0.772 |
| *Sex* |  |  |  |  |
| Female | 233 (25.9) | 60 (16.3) | 132 (20.2) | <0.001 |
| Male MSM <sup>a</sup> | 459 (50.8) | 253 (70.0) | 419 (63.4) |  |
| Male non-MSM | 203 (22.6) | 49 (13.4) | 110 (16.2) |  |
| Missing | 5 (0.55) | 5 (1) | 2 (0.3) |  |
| ***Race*** |  |  |  |  |
| White | 478 (53.1) | 252 (68.7) | 442 (66.6) | < 0.001 |
| Black | 263 (29.3) | 55 (14.9) | 130 (19.6) |  |
| Other | 159 (17.6) | 60 (16.3) | 91 (13.7) |  |
| ***Region*** |  |  |  |  |
| Eastern Ontario | 127 (14.1) | 24 (6.5) | 22 (3.3) | < 0.001 |
| Northern Ontario | 18 (2) | 5 (1.4) | 2 (0.3) |  |
| Southwestern Ontario | 99 (11) | 62 (16.9) | 174 (26.2) |  |
| Toronto | 656 (72.8) | 276 (75.2) | 465 (70.1) |  |
| ***Education*** |  |  |  |  |
| Elementary | 25 (2.7) | 10 (2.7) | 14 (2.1) | 0.246 |
| High school | 239 (26.5) | 126 (34.3) | 224 (33.7) |  |
| College | 210 (23.3) | 64 (17.4) | 139 (20.9) |  |
| Higher education | 363 (40.3) | 162 (44.1) | 284 (42.8) |  |
| Missing | 63 (7) | 5 (1.4) | 2 (0.3) |  |
| ***Employment status*** |  |  |  |  |
| Employed | 433 (48.1) | 182 (49.6) | 327 (49.3) | 0.285 |
| Unemployed | 463 (51.4) | 182 (49.6) | 336 (50.7) |  |
| Missing | 4 (0.4) | 3 (0.8) | 0 (0) |  |
| ***Gross annual income*** |  |  |  |  |
| <50,000 | 374 (41.5) | 144 (39.2) | 280 (42.2) | 0.287 |
| < 50,000-70,000 | 103(11.4) | 45 (12.2) | 75 (11.3) |  |
| < 70,000-80,000 | 121 (13.4) | 41 (11.1) | 76 (11.4) |  |
| >100,000 | 173 (19.2) | 94 (25.6) | 150 (22.6) |  |
| Missing | 129 (14.3) | 43 (11.7) | 82 (12.3) |  |
| ***Stable relationship*** |  |  |  |  |
| Yes | 376 (41.6) | 154 (42.0) | 246 (37.1) | 0.17 |
| Unstable relationship | 521 (57.8) | 212 (57.8) | 417 (62.9) |  |
| Missing | 3 (0.3) | 1 (0.3) | 0 (0) |  |
| ***Adherence to ART*** |  |  |  |  |
| < 95% | 295 (32.7) | 137 (37.3) | 247 (37.3) | 0.014 |
| ≥ 95% | 600 (66.6) | 225 (61.3) | 412 (62.1) |  |
| Missing | 5 (0.5) | 5 (1.3) | 4 (0.6) |  |
| ***Mental health conditions*** |  |  |  |  |
| Depression | 197 (21.8) | 44 (12.0) | 167 (25.1) | 0.190 |
| Alcohol use disorder<br>syndrome <sup>d</sup> | 67 (7.4) | 34 (9.2) | 65 (9.8) |  |
| No | 824 (91.6) | 331 (90.1) | 598 (90.1) | 0.248 |
| Yes |  |  |  |  |
| ***Stigma*** |  |  |  |  |
| Stigmatized | 125 (13.8) | 12 (3.28) | 92 (13.8) | 0.5083 |
| Not stigmatized | 8 (0.88) | 2 (0.54) | 7 (1.05) |  |
| Missing | 767 (85.2) | 353 (96.1) | 564 (85.0) |  |
| ***Quality of life*** |  |  |  |  |
| MCS | 50.2 [41.25;<br>57.00] | 49.2 [40.82;55.32] | 50.9 [38.67;56.30] | 0.403 |
| PCS | 53.1 [43.8;57.2] | 53.5 [44.87;57.35] | 52.7 [43.57;56.70] | 0.548 |
| ***Viral load*** |  |  |  |  |
| ≤ 40 | 766 (85.0) | 336 (91.6) | 579 (87.3) | 0.02 |
| > 40 | 104 (11.5) | 26 (7.0) | 70 (10.5) |  |
| Missing | 30 (0.5) | 5 (1.4) | 14 (2.1) |  |
| <sup>a</sup> Men who report having sex with other men<br><sup>b</sup> Immigrant > 10 years<br><sup>c</sup> Immigrant <10 years<br><sup>d</sup> Alcohol Use Disorder Identification Test (AUDIT-10) |  |  |  |  |

For demographic variables, we combined sex (female versus male) and sexual orientation (men who have sex with men (MSM) vs non-MSM) to derive a single variable named sex, compromising three categories: females, male MSM, and male non-MSM and relationship status categorized as stable (married, living common-law, living in a committed relationship) vs unstable relationship (widowed, separated/divorced, single).

Clinical variables were extracted on the following health conditions: alcohol use disorder syndrome use was defined as on the Alcohol Use Disorder Identification Test (AUDIT-10) with harmful alcohol measured from 10-items (a score of ≥8 regardless of gender/sex) [25], depression measured by the Patient Health Questionnaire (PHQ) scale with nine items [26], smoking cigarettes/tobacco defined as heavy smokers versus light smokers (heavy smokers defined as people who have been smoking at least 20 cigarettes daily) [27]; and diagnosis of mental health comorbidities was based on self-report in the OCS question.

We used a revised 10-item HIV-related stigma scale categorized into four major components of HIV-related stigma: personalized stigma, worries about disclosure of status, negative self-image, and sensitivity to public reactions about HIV status. Individuals who responded with “agree’ and “strongly agree” were identified as experiencing stigma in at least one of the four components [28].

#### Sample size calculations

The study was planned with a sample size of 1,930 subjects, providing 80% power to detect a difference of 20% or greater between suboptimal and optimal adherence to ART between the groups. [29]. The Type I error probability associated with the test of the null hypothesis is 0.05 for the two-tailed chi-squared statistic. (PS: Power and Sample Size Calculation version 3.1.2, 2014 by W.D. Dupont & W.D. Plummer Jr).

### Data analysis

Statistical analysis was performed using R software version 4.4.1. We used descriptive statistics to analyze participants’ characteristics, reporting proportions for categorical variables and median with interquartile range (IQR) for continuous variables. The latter were compared using the Wilcoxon rank-sum test, as these scores are not normally distributed [30]. Chi-square tests were used for categorical variables.

The variable selection was guided by a priori knowledge of their association with adherence to ART, considering collinearity between variables and potential confounding. Age, sex, ethnicity, employment, education level, substance use, stigma and living alone were included in all models as potential confounders, regardless of their significance [3, 11, 31, 32]. To select the remaining variables for our models, we used the ‘majority selection method’ and selected the variables that appeared in at least half of the models [33]

#### Regression Analysis

We conducted a logistic regression analysis for dichotomous outcomes (adherence to ART and suppression of the viral load) and a multiple linear regression for continuous outcomes (quality of life). To check the model fit for multiple linear regression models, we used a pooled R^2^. The dichotomous outcomes are reported as odds ratio (OR) and 95% confidence intervals, and continuous outcomes are reported as mean differences with 95% confidence intervals. The statistically significant level is set when the *p*-value is <0.05 or the 95% CI excludes the null value.

The OCS questionnaire had data missing at random (MAR), so we performed ten imputations for each model and combined the results using Rubin’s rules [33, 34].

#### Subgroup analysis

We performed subgroup analysis to assess the differences in health outcomes (adherence to ART, quality of life and viral load) of people living with HIV from different socio-demographics in Ontario, Canada. The data from the OCS Questionnaire 2022 was collected during the COVID-19 pandemic when virtual visits were first introduced. [15]. Since government COVID-19 policies and users’ preferences may have impacted decisions to attend virtual visits, we compared outcomes during and after lockdowns [35].

## Results

In 2022, the OCS questionnaire was completed by 2155 people, with 1930 providing details on the type of care they received (S 1 Fig). That year, 1021/1930 (53%) HIV care visits were conducted by telephone, 23/1930 (1.2%) via computer visits, and 1563/1930 (80%) were provided through in-person visits. The median age of the participants was 55 years [IQR: 45-62]. In 2022, stratum-specific proportions of participants in the three different types of care modalities (i.e. in-person, virtual and participants who used both in-person and virtual care) varied by participant characteristics (Table 1). Notably,1493/1930 (78%) participants were men, with the majority being MSM, compromising 58.6% (1131/1930). Regarding care modality preferences, in-person visits were most preferred across all genders. Table 1 provides the statistical relationship between all other variables and the type of care.

**Fig 1.**
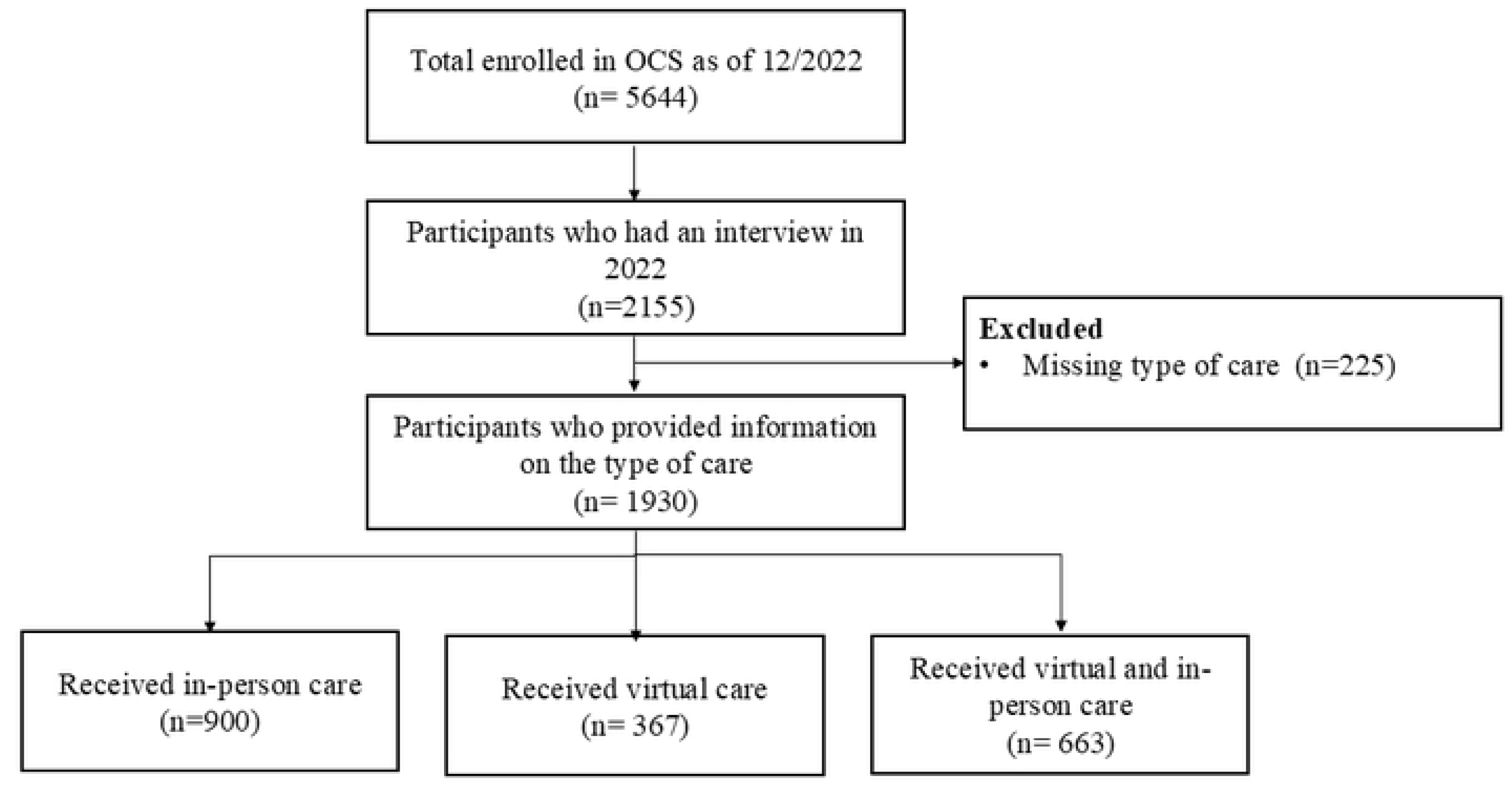
Study flow diagram of participants who received HIV care in 2022.

### Adherence to ART

There were 600 (66.6%) participants with optimal ART adherence. According to our logistic regression analysis, the odds of adherence to ART were higher for participants who used virtual care than in-person care in both adjusted and unadjusted analysis (OR 1.47 %, 95% CI 1.14, 1.89; AOR 1.31%, 95% CI 1.00-1.71).

### Viral load

There were 766 (85.0%) participants with adequate viral load suppression. The odds of viral load suppression were higher in the virtual group than in person-care (OR 1.81, 95%CI: 0.69-2.85; AOR 1.67, 95% CI:1.03-2.63).

### Quality of life

The average MCS score was 50.2 and, the average PCS score was 53.1. According to our adjusted multiple regression model, combined virtual and in-person care is associated with an improved quality of life in terms of MCS compared to in-person care (Mean difference (MD) -0.90, 95 % CI - 2.10,0.30; adjusted MD 0.960, 95% CI 0.052,1.869). All results can be found in Table 2.

**Table 2:** Multivariable regression analysis of HIV-related outcomes of people receiving care in Ontario, Canada.

| Variable | Unadjusted Effect<br>Estimates (95%CI) | p-value | Adjusted Effect<br>Estimates (95%<br>CI) | p-value |
| --- | --- | --- | --- | --- |
| ***Adherence to ART (OR)*** |  |  |  |  |
| In-person(ref) | 1.00 (ref) |  | 1.00 (ref) |  |
| Virtual and in-person | 1.23 (0.99, 1.52) | 0.05 | 1.03 (0.82, 1.30) | 0.743 |
| Virtual | 1.47 (1.14, 1.89) * | 0.003 | 1.30 (1.00, 1.70) * | 0.048 |
| ***Viral load (OR) <40 ml/cc preferred*** |  |  |  |  |
| In-person(ref) | 1.00 (ref) |  | 1.00 (ref) |  |
| Virtual and in-person care | 1.13 (0.52, 1.56) | 0.431 | 1.08 (0.76, 1.53) | 0.651 |
| Virtual care | 1.81 (0.69, 2.85) | 0.010 | 1.67 (1.03, 2.63) | 0.0374 |
| ***<sup>1</sup>MCS*** |  |  |  |  |
| In-person visit | 0 (ref) |  | 0 (ref) |  |
| Virtual and in-person | -0.90 (-2.10,0.30) | 0.142 | 0.96 (0.05,1.86) * | 0.038 |
| Virtual | -0.53 (-1.99, 0.92) | 0.469 | 0.13 (-0.94, 1.2) | 0.810 |
| ***<sup>2</sup>PCS*** |  |  |  |  |
| In-person visit | 0 (ref) |  | 0 (ref) |  |
| Virtual and in-person | -0.90 (-2.10, 0.30) | 0.142 | -0.91 (-1.24,0.85) | 0.711 |
| Virtual | -0.53 (-1.99, 0.92) | 0.469 | 0.09 (-1.14,1.32) | 0.811 |
| <sup>a</sup> Adjusted for covariates<br>* Statistically significant<br><sup>1</sup> Mental Component Summary Score<br><sup>2</sup> Physical Component Summary Score |  |  |  |  |

### Sub-group analysis

Among patients whose physicians based their preference of the type of care mode on the viral load, receiving in-person care was associated with lower odds of adherence to ART than receiving virtual care (OR 0.29, 95% CI 0.10-0.83; AOR 0.28, 95% CI 0.09-0.86).

Among patients whose physicians based their preference of the type of care mode on the viral load, the receipt of both virtual and in-person care was associated with decreased MCS quality of life compared with participants who used the in-person care mode. (MD -5.60, 95% CI -9.46 - 1.75; Adjusted MD -3.75, 95% CI -6.51—0.99). Detailed findings are provided in S 1 Table.

## Discussion

This study examined three types of care used in the OCS cohort 2022: in-person, virtual and patients using both. Patients across all demographic and medical backgrounds used virtual care options. In this cohort, the participants who used virtual care mode most frequently were 51-60, perhaps suggesting that older patients are comfortable with technology. However, the predominant mode of care within this cohort remained in-person visits. In this cohort consisting of participants living with HIV in Ontario, we observed that participants with better adherence to ART and with viral load suppression preferred the virtual care, and participants with a higher MCS quality of life utilized both virtual and in-person care.

Sex was the strongest predictor of the type of care used, with male MSM favouring in-person care. Participants residing in Toronto preferred virtual care compared to those in Eastern Ontario, though these groups overlap, as many male MSM in Toronto might be the same individuals. Participants with any form of depression tended to use both virtual and in-person care, possibly choosing their care mode based on the severity of their depression at the time of the visit.

Our subgroup analysis based on physicians’ preference for the type of visit by viral load revealed decreased ART adherence within the virtual care group. However, the primary analysis showed a 30% improvement in ART adherence among participants using virtual visits compared to in-person groups. This suggests physicians preferred in-person visits for patients with unsuppressed viral lows and poor ART adherence.

This study has several limitations that prevent drawing definitive conclusions. The cross-sectional design prevents us from establishing causality, making it difficult to determine whether virtual care directly influences HIV related-health outcomes. Additionally, the data analyzed is from 2022, a period when virtual care was newly introduced and still in the early stages of implementation, including the development of specific standards, equipment selection and user training [38, 49]. Moreover, during the study period, decisions regarding the type of care were often influenced by the government’s lockdown orders rather than the preferences of physicians or patients [38]. Non-response bias further restricts the conclusion of differences between care modes. Furthermore, challenges related to optimizing virtual care usability and user-friendliness remain unexplored.

The OCS employs a standardized questionnaire developed in consultation with the OHTN, CAB and the governance committees. However, the primary outcome—ART adherence—is based on self-reporting, which is subjective and prone to recall bias [3].

The study’s findings are influenced by selection and information biases, as participants are predominantly aged 51-55, with high incomes and stable housing. This limits the understanding of age-related acceptance and barriers. The cohort mainly includes highly engaged individuals, which doesn’t fully represent overall care engagement. Younger, tech-savvy individuals and rural populations are underrepresented in this cohort, limiting insights into the broader applicability of virtual care and rural-urban differences [32, 35].

Despite potential sample biases, the study’s large sample size and real-time data collection enhance the robustness of its findings.

The study was guided by a CAB, whose input was integrated throughout the study, including their feedback on result interpretation. Collaborating closely with experienced HIV care physicians provides valuable insights into clinical practice dynamics and potential implementation strategies based on study outcomes [36].

### Future research

It’s essential to recognize that this research is exploratory, and the COVID-19 pandemic lockdowns confound it. Future research should explore the impact of virtual visits on HIV care and improve access. Key issues include standards, licensure, equity, and payment systems. Virtual visits can help address physician shortages, especially in rural areas, by offering flexible, quality care. Expanding virtual care to all socioeconomic groups is vital. Redesigning care based on necessity and feasibility could enhance access and outcomes [37, 38].Future studies should assess system-wide effects, user satisfaction, cost-effectiveness, and physician views while linking OCS data with ICES data to analyze clinical outcomes and patient preferences.

## Conclusion

In our study in the OCS 2022 of people living with HIV, participants with better self-reported adherence to ART and a suppressed viral load preferred virtual care as compared to in-person care. Moreover, participants who used both virtual and in-person care reported higher mental health quality of life.

## Data Statement

The data is obtained from OHTN from the OCS study. It is unethical for the authors to share the data set publicly. However, the data request can be made directly with OHTN by submitting a [Research Application Process (RAP)](https://ohtncohortstudy.ca/research/).

## Authors’ contributions

A.J., L.M., and N.R. contributed to the study’s concept. The study design was finalized after feedback from all the authors (L.M., N.R., M.G., D.M., and A.J.). OHTN provided the final approval and the data. N.R. and A.J. conducted the analysis. N.R. prepared the original draft. All authors critically revised, approved and agreed to be held accountable for all aspects of this work. Realize provided support and guidance for CAB recruitment, consultation on community engagement and study design.

## Acknowledgements

The OHTN Cohort Study Team consists of Dr. Ann Burchell (Interim Principal Investigator), St. Michael’s Hospital and University of Toronto; Dr. Anita Benoit (Co-investigator), University of Toronto; Dr. Lawrence Mbuagbaw (Co-Investigator), McMaster University; Dr. Sergio Rueda, CAMH and University of Toronto; Dr. Gordon Arbess, St. Michael’s Hospital; Dr. Jeffrey Cohen, Windsor Regional Hospital; Dr. Curtis Cooper, Ottawa General Hospital; Dr. Maheen Saeed, University of Ottawa Health Services; Dr. Mona Loutfy, Maple Leaf Medical Clinic; Dr. David Knox, Maple Leaf Medical Clinic; Dr. Nisha Andany, Sunnybrook Health Sciences Centre; Dr. Sharon Walmsley, Toronto General Hospital; Dr. Michael Silverman, St. Joseph’s Health Care; Ms. Tammy Bourque, Health Sciences North; Wangari Tharao, Women’s Health in Women’s Hands Community Health Centre; Holly Gauvin, Elevate NWO; Dr. Jeffrey Craig, Lakeridge Health, Dr. Jorge Martinez-Cajas, Kingston Health Sciences Centre, and Dr. Marek Smijea, Hamilton Health Sciences Centre.

We gratefully acknowledge all of the people living with HIV who volunteered to participate in the OHTN Cohort Study and the work and support of the OCS Governance Committee: (Dane Record, Jasmine Cotnam, Barry Adam, Adrian Betts, Cornel Gray, Ruth Cameron, Rodney Rousseau, Mary Ndung’u, Viviana Santibañez, YY Chen, Jason Brophy, Aaron Bowerman), OCS Scientific Steering Committee (Dr. Sergio Rueda, Dr. Barry Adam, Dr. David Brennan, Dr. Trevor Hart, Dr. Lance McReady, Dr. Lawrence Mbuagbaw, Dr. Anita Benoit, Dr. Mona Loutfy, Dr. Curtis Cooper, Dr. Sean Hillier, Mr. Pierre Giguere, Dr. Kelly O’Brien).

We thank all the interviewers, data collectors, research associates and coordinators, nurses and physicians who provide support for data collection and extraction. The authors wish to thank the OCS staff for data management, IT support, and study coordination: Lydia Makoroka, Lucia Light, Nahid Qureshi, Namita Prabhu, Tsegaye Bekele, and Mustafa Karacam.

We also acknowledge the Public Health Laboratories, Public Health Ontario, for supporting record linkage with the HIV viral load database.

The OHTN Cohort Study is supported by the Ontario Ministry of Health and Long-Term Care.

The opinions, results and conclusions are those of the authors and no endorsement by the Ontario HIV Treatment Network or Public Health Ontario is intended or should be inferred.

## Supporting information

**S 1 Fig.** Study flow diagram of participants who received HIV care in 2022.

**S 1 Table.** Multivariable subgroup regression analysis of HIV-related outcomes of people receiving care in Ontario, Canada

## References

1. The Ontario HIV Epidemiology and Surveillance Initiative. Current Trend for HIV in Ontario: OHESI; 2023 [Available from: https://www.ohesi.ca].

2. Wilton J, Liu J, Sullivan A, Rachlis B, Marchand-Austin A, Giles M, et al. Trends in HIV care cascade engagement among diagnosed people living with HIV in Ontario, Canada: A retrospective, population-based cohort study. PLoS One. 2019;14(1):e0210096.

3. Rachlis B, Burchell AN, Gardner S, Light L, Raboud J, Antoniou T, et al. Social determinants of health and retention in HIV care in a clinical cohort in Ontario, Canada. AIDS Care. 2017;29(7):828–37.

4. Abebe Moges N, Olubukola A, Micheal O, Berhane Y. HIV patients retention and attrition in care and their determinants in Ethiopia: a systematic review and meta-analysis. BMC Infect Dis. 2020;20(1):439.

5. Benoit AC, Burchell AN, O’Brien KK, Raboud J, Gardner S, Light L, et al. Examining the association between stress and antiretroviral therapy adherence among women living with HIV in Toronto, Ontario. HIV Res Clin Pract. 2020;21(2-3):45–55.

6. Mbuagbaw L, Sivaramalingam B, Navarro T, Hobson N, Keepanasseril A, Wilczynski NJ, et al. Interventions for enhancing adherence to antiretroviral therapy (ART): a systematic review of high quality studies. AIDS Patient Care and STDs. 2015;29(5):248–66.

7. Mbuagbaw L, Hajizadeh A, Wang A, Mertz D, Lawson DO, Smieja M, et al. Overview of systematic reviews on strategies to improve treatment initiation, adherence to antiretroviral therapy and retention in care for people living with HIV: part 1. BMJ Open. 2020;10(9):e034793.

8. Yehia BR, Stephens-Shields AJ, Fleishman JA, Berry SA, Agwu AL, Metlay JP, et al. The HIV Care Continuum: Changes over Time in Retention in Care and Viral Suppression. PLOS One. 2015;10(6):e0129376.

9. Kay ES, Batey DS, Mugavero MJ. The HIV treatment cascade and care continuum: updates, goals, and recommendations for the future. AIDS Res Ther. 2016;13:35.

10. Government of Ontario. Virtual Care 1: Comprehensive and Limited Virtual Care Services 2022 [cited 2023 24th August 2023]. Available from: https://www.ontario.ca/document/education-and-prevention-committee-billing-briefs/virtual-care-1-comprehensive-and-limited#section-2.

11. Heyworth L, Kirsh S, Zulman D, Ferguson JM, Kizer KW. Expanding access through virtual care: The VA’s early experience with Covid-19. NEJM Catalyst Innovations in Care Delivery. 2020.

12. Government of Canada. Pan-Canadian virtual care priorities in response to COVID-19 2021.

13. Gomez T, Anaya YB, Shih KJ, Tarn DM. A Qualitative Study of Primary Care Physicians’ Experiences With Telemedicine During COVID-19. The Journal of the American Board of Family Medicine. 2021;34(Supplement):S61–S70.

14. Glauser W. Virtual care is here to stay, but major challenges remain: CMAJ. Canadian Medical Association Journal. 2020;192(30):E868–E9.

15. Government of Ontario. OHIP Info Bulletins. 2022.

16. Budak JZ, Scott JD, Dhanireddy S, Wood BR. The Impact of COVID-19 on HIV Care Provided via Telemedicine-Past, Present, and Future. Curr HIV/AIDS Rep. 2021;18(2):98–104.

17. McGinnis KA, Skanderson M, Justice AC, Akgun KM, Tate JP, King JT, Jr., et al. HIV care using differentiated service delivery during the COVID-19 pandemic: a nationwide cohort study in the US Department of Veterans Affairs. J Int AIDS Soc. 2021;24 Suppl 6:e25810.

18. Dorn SD. Backslide or forward progress? Virtual care at U.S. healthcare systems beyond the COVID-19 pandemic. NPJ Digit Med. 2021;4(1):6.

19. The Ontario HIV Treatment Network. HIV in Ontario 2023 [Available from: https://www.ohtn.on.ca.

20. Ontario HIV treatment Network OCS (OHTN Cohort Study). Research Policies [Available from: https://ohtncohortstudy.ca/wp-content/uploads/2022/12/OCS-Research-Policies_6-Oct-2022_Clean.pdf.

21. Rourke SB, Gardner S, Burchell AN, Raboud J, Rueda S, Bayoumi AM, et al. Cohort profile: the Ontario HIV Treatment Network Cohort Study (OCS). Int J Epidemiol. 2013;42(2):402–11.

22. Lo Hog Tian JM, Watson JR, Deyman M, Tran B, Kerber P, Nanami K, et al. Building capacity in quantitative research and data storytelling to enhance knowledge translation: a training curriculum for peer researchers. Research Involvement and Engagement. 2022;8(1):69.

23. Cashman SB, Adeky S, Allen III AJ, Corburn J, Israel BA, Montaño J, et al. The power and the promise: working with communities to analyze data, interpret findings, and get to outcomes. American Journal of Public Health. 2008;98(8):1407–17.

24. Ding X, Abner EL, Schmitt FA, Crowley J, Goodman P, Kryscio RJ. Mental Component Score (MCS) from Health-Related Quality of Life Predicts Incidence of Dementia in U.S. Males. J Prev Alzheimers Dis. 2021;8(2):169–74.

25. Bush K, Kivlahan DR, McDonell MB, Fihn SD, Bradley KA. The AUDIT alcohol consumption questions (AUDIT-C): an effective brief screening test for problem drinking. Ambulatory Care Quality Improvement Project (ACQUIP). Alcohol Use Disorders Identification Test. Arch Intern Med. 1998;158(16):1789–95.

26. Löwe B, Kroenke K, Herzog W, Gräfe K. Measuring depression outcome with a brief self-report instrument: sensitivity to change of the Patient Health Questionnaire (PHQ-9). Journal of Affective Disorders. 2004;81(1):61–6.

27. Machado RC, Vargas HO, Baracat MM, Urbano MR, Verri Jr WA, Porcu M, et al. N-acetylcysteine as an adjunctive treatment for smoking cessation: a randomized clinical trial. Brazilian Journal of Psychiatry. 2020;42:519–26.

28. Etowa J, Hannan J, Babatunde S, Etowa EB, Mkandawire P, Phillips JC. HIV-related stigma among black mothers in two north American and one African cities. Journal of Racial and Ethnic Health Disparities. 2020;7:1130–9.

29. Cote J, Godin G, Ramirez-Garcia P, Rouleau G, Bourbonnais A, Gueheneuc YG, et al. Virtual intervention to support self-management of antiretroviral therapy among people living with HIV. J Med Internet Res. 2015;17(1):e6.

30. Bridge PD, Sawilowsky SS. Increasing physicians’ awareness of the impact of statistics on research outcomes: comparative power of the t-test and Wilcoxon rank-sum test in small samples applied research. Journal of Clinical Epidemiology. 1999;52(3):229–35.

31. Logie C, James L, Tharao W, Loutfy M. Associations between HIV-related stigma, racial discrimination, gender discrimination, and depression among HIV-positive African, Caribbean, and Black women in Ontario, Canada. AIDS patient care and STDs. 2013;27(2):114–22.

32. Burchell AN, Gardner S, Light L, Ellis BM, Antoniou T, Bacon J, et al. Implementation and operational research: engagement in HIV care among persons enrolled in a clinical HIV cohort in Ontario, Canada, 2001–2011. JAIDS Journal of Acquired Immune Deficiency Syndromes. 2015;70(1):e10-e9.

33. Heymans MaE I. Applied missing data analysis with SPSS and (R) Studio. Heymans and Eekhout;2019.

34. Van Buuren S. Flexible imputation of missing data: CRC press; 2018.

35. Ferguson JM, Jacobs J, Yefimova M, Greene L, Heyworth L, Zulman DM. Virtual care expansion in the Veterans Health Administration during the COVID-19 pandemic: clinical services and patient characteristics associated with utilization. J Am Med Inform Assoc. 2021;28(3):453–62.

36. Realize. Fostering positive change for people living with HIV and other episodic disabilities.. 2023 [Available from: http://forum.realizecanada.org/about-us/.

37. Lawal FJ, Omotayo MO, Lee TJ, Srinivasa Rao ASR, Vazquez JA. HIV Treatment Outcomes in Rural Georgia Using Telemedicine. Open Forum Infect Dis. 2021;8(6):ofab234.

38. León A, Cáceres C, Fernández E, Chausa P, Martin M, Codina C, et al. A new multidisciplinary home care telemedicine system to monitor stable chronic human immunodeficiency virus-infected patients: a randomized study. PLoS One. 2011;6(1):e14515.

